# Electroencephalography Markers of Perinatal Depression and Anxiety in a Diverse Longitudinal Cohort

**DOI:** 10.1101/2024.10.23.24315990

**Authors:** Maigh Attre, Suzanne Alvernaz, Elizabeth Wenzel, Abigail Pettineo, Sarah Parikh, Catherine Conway, Molly Kroeger, Mary Kimmel, Olu Ajilore, Pauline Maki, Alex Leow, Beatriz Peñalver Bernabé

**Affiliations:** Richard and Loan Hill Department of Biomedical Engineering, Colleges of Engineering and Medicine, University of Illinois Chicago, Chicago, IL, USA; Department of Psychology, Colleges of Liberal Arts and Science, University of Illinois Chicago, Chicago, IL, USA; Department of Psychiatry and Neurobehavioral Sciences, College of Medicine, University of Virginia, Charlottesville, VA; Department of Psychiatry, College of Medicine, University of Illinois Chicago, Chicago, IL, USA; Department of Psychiatry, College of Medicine, Washington University, St. Louis, MO, USA; Department of Obstetrics and Gynecology, College of Medicine, University of Illinois Chicago, Chicago, IL, USA; Center of Bioinformatics and Computational Biology, Chicago, IL, USA

**Keywords:** Electroencephalography, perinatal depression, perinatal anxiety

## Abstract

Mental health disorders, such as depression and anxiety, are highly prevalent during pregnancy and postpartum and can have severe consequences on maternal morbidity and mortality, and on infant neurodevelopment. Yet the neurobiological mechanisms of these disorders are underexplored. Critically absent are longitudinal studies of mental health and brain alterations across the entire perinatal period using electroencephalography (EEG). In this study, a racially, ethnically, and socioeconomically diverse perinatal cohort (n=61, 45% Non-Hispanic Black, 32% Hispanic, 65% public insurance) was followed bimonthly from early pregnancy through postpartum. At each visit, participants completed an 8-minute closed-eye resting state EEG recording and self-reported validated questionnaires to assess depression and anxiety symptoms. Linear regression models showed EEG-measured relative delta waves were negatively associated with both depression and anxiety severity in the first trimester and postpartum. Alpha waves were positively associated with elevated symptoms of anxiety at postpartum. Early pregnancy alpha and delta waves were also associated with late third trimester pregnancy and postpartum depression and anxiety. This study is the first to report that specific brain waves are associated with mood and anxiety disorders during perinatal periods characterized by large reproductive hormonal fluctuations. This work is an initial step to understanding how changes in dynamic brain activity during the perinatal period may be used as possible unbiased markers of perinatal mood and anxiety disorders.

## INTRODUCTION

Perinatal mental health disorders such as perinatal depression and anxiety, are prevalent (10-20%) (1,2), can be detrimental to maternal health, and could contribute to suicidality which accounts to over 20% of deaths within the first year postpartum (3). Depression and anxiety symptomatology, even levels below the diagnostic criteria for perinatal depression and anxiety, can have adverse effects to maternal health (4,5). Black and Hispanic perinatal mental health has been especially under researched even though pregnant women of color being disproportionally affected by perinatal mood disorders (6–10). Despite the high prevalence of mental health disorders during the perinatal period, identifying underlying neurologic alterations is challenging due to the complex physiological adaptations of pregnancy. During pregnancy, hormones like estradiol and progesterone increase substantially, followed by a drastic withdrawal after childbirth. These fluctuations are correlated with structural brain modifications during gestation and postpartum (11–17). Emerging work has linked both perinatal depression and anxiety to distinct temporal trajectories in pregnancy hormones like progesterone and allopregnanolone levels across the perinatal period (18–21). While systemic hormonal and brain structural adaptations are linked in normal pregnancy(22), the functional characterization of brain activity changes is limited across gestation especially in the context of perinatal depression and anxiety. Brain functionality markers of mood disorders would be highly clinically relevant for perinatal health.

Electroencephalography (EEG) is a non-invasive tool to measure synchronous neuronal electrical activity. It has been shown to be sensitive to mental health disorders, including those that occur during periods of profound hormonal fluctuation such as the menstrual cycle or menopause (23,24). Many EEG studies have examined mental health in association with brain electrical activity states classified by brain wave frequency bands: delta waves (1 - 4 Hz) are associated with deep sleep; theta waves (4 - 8 Hz) with emotional processing; alpha waves (8 - 13 Hz) with inactivity in the brain and relaxation; beta waves (13 - 32 Hz) with concentration; and gamma waves (32-60 Hz) with attention and sensation (25). These waves have been implicated in general depression (25,26), anxiety (27), and other mood disorders (28) outside the perinatal period. Specifically, in non-pregnant cohorts, increased asymmetry in frontal alpha waves and increased theta waves were associated with major depressive disorder (25,26), whereas increased alpha, theta, and beta waves were associated with generalized anxiety disorder (27,29,30). During the menstrual cycle, several EEG studies have reported differences in alpha symmetry in menstruation related mood disorders during periods of hormonal sensitivity, specifically during the progesterone and estradiol surge of the luteal phase(23). This implies hormones may mediate neural activity in other reproductive stage mental health disorders. During menopause, EEGs of depressed individuals demonstrate similar asymmetric alterations, lower relative alpha power, and higher delta power compared to non-depressed menopausal controls (24). These findings suggest that physiologic hormonal fluctuations throughout reproductive life, such as progesterone and estrogen shifts, influence neural electrical activity. Thus, EEGs could provide possible markers of mood disorders during these periods. However, the potential association between EEG characteristics of the perinatal brain and mood disorders across gestation and postpartum remains almost unknown.

EEG studies of depression and anxiety are limited in the perinatal period. While some EEG studies of uncomplicated pregnancy have examined pregnancy related sleep disturbance (31), perinatal associations between maternal-offspring right frontal activation (32), general activity across the third trimester and postpartum (14,33), and working memory across gestation (34), very few EEG studies have examined perinatal mood and anxiety symptoms. Importantly, those pioneer studies are cross-sectional and has been performed mostly in the third trimester, rather than longitudinally, from early pregnancy through postpartum and thus not capturing the periods of profound physiologic and hormonal change that occur in early pregnancy or at birth. Peng and colleagues identified decreases in EEG measured brain network connectivity in individuals reporting elevated levels of depression in a cohort of 62 women in the late third trimester (35). Levy and colleagues identified decreased beta oscillations in relation to anxiety during the third trimester in a large cohort of 135 pregnant women(36). To fully characterize the dynamics of mental health across the perinatal window, studies that assess individual alterations from early pregnancy through postpartum are needed. Furthermore, none of the perinatal EEG studies to date have focused on minority populations, even though Black and Hispanic women are less likely to be screened and treated for perinatal mental health disorders (10). In a longitudinal study of perinatal depression and anxiety in a diverse cohort of women, we found that depression and anxiety symptoms were generally most severe in the first trimester, especially for Hispanic women, underscoring the importance of studying mental health early in pregnancy (6). Yet, Black and Hispanic women are underrepresented in maternal EEG studies (37). Increasing the racial and ethnic diversity in EEG studies of perinatal mental health is essential given the increased prevalence of these disorders in women of color, and their effect on maternal and infant wellbeing.

Here, we aim to address these gaps by conducting a longitudinal, resting-state EEG study in a diverse perinatal cohort with the goal of identifying alterations in brain electrophysiology across pregnancy and postpartum in relation to depression and anxiety symptoms. We used a commercially available portable EEG (pEEG), a relatively low-cost, non-invasive method that is easy to use in clinical settings. We hypothesized that brain electrical activity would be associated with the severity of perinatal depression and anxiety across the perinatal period, and that these associations would be most evident during rapid changes in pregnancy hormones, such as early in pregnancy and at postpartum.

## METHODS

### Study Design

This work is a sub-study of an ongoing observational clinical study Moms and Mental Health (MoMent) at the University of Illinois Chicago (IRB #2014-2035) (6,38–40). The parent study is investigating longitudinal alterations in the microbiota-gut-brain axis in relation to mental health during the perinatal period. In this observational study, participants answered a battery of questionnaires about socio-demographic, prior medical history and mental health, and provided multiple biological samples. A subset of these participants also underwent assessments of brain activity using a commercial pEEG. Data used for this work was collected between January, 2022 and September, 2024. All study participants provided informed consent prior to data collection. Inclusion criteria: being English speaking, over 18 years of age, and less than 16 gestational weeks (GWs) pregnant. Exclusion criteria: pregnancies with more than one fetus; use of *in-vitro* fertilization for current pregnancy; use of oral antibiotics, antifungals or antivirals or laxatives for weight loss in the last 3 months; active diagnosis of HIV or cancer; history of gastrointestinal surgeries; eating disorder or chronic diarrhea diagnosis within the last 6 months; or history of substance abuse within the last 3 months. Study visits were completed at approximately less than 16 GWs, 18-22 GWs, 24-28 GWs, 32-36 GWs and 6-12 weeks postpartum (**Figure 1**).

**Figure 1:**
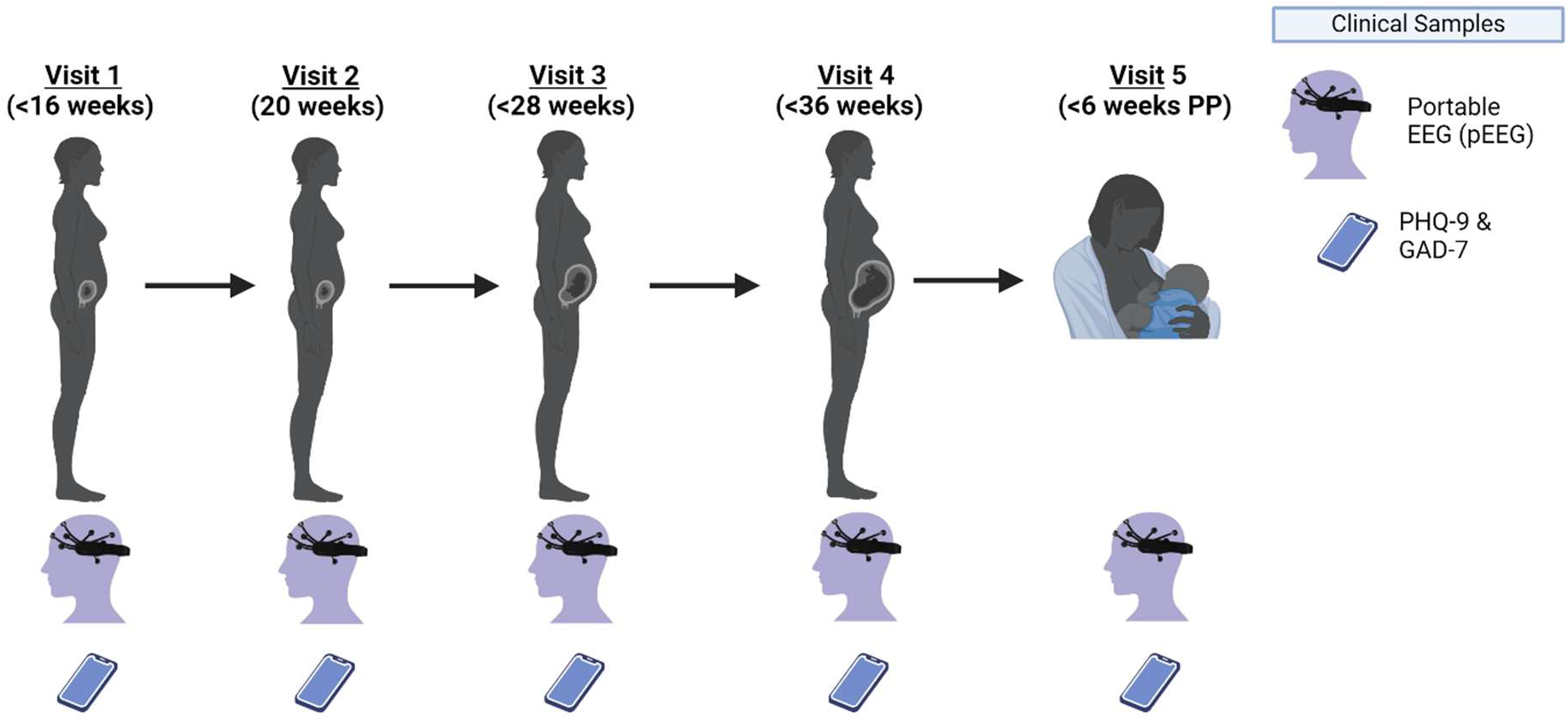
Study Design. Study participants completed up to five EEG recordings and self-reported mental health questionnaires. GAD-7: the general anxiety disorder-7; PHQ-9: the patient health questionnaire-9 (PHQ-9), PP: postpartum. Created in Biorender.

### Mental Health Assessments

Study participants electronically completed the patient health questionnaire-9 (PHQ-9)(41,42), which assessed depression symptoms, and general anxiety disorder-7 (GAD-7)(43,44), which assessed anxiety symptoms, within 24 hours of each brain assessment. We employed the following cutoffs to define both depression and anxiety severity: normal (0-4), mild (5-9), and moderate/severe (≥10).

### Electroencephalographic assessments of brain electrical activity

Participants completed an 8-minute closed-eye resting state EEG recording at each visit using the EPOC X 14 Channel Wireless EEG Headset from EMOTIV (45). Felt pads for each electrode were soaked in standard contact lens saline solution (Bausch + Lomb Biotrue Contact Lens Solution). Participants were asked to remove any glasses, hats, or headbands. The portable EEG (pEEG) device was fit around participant’s hair following the manufacturer instructions. If a study participant was wearing a wig, the device was placed on top as normal and documented in the recording notes. Readings were completed in a quiet room with no lights. Participants were instructed to sit as still as possible with their eyes closed and let their mind wander for the duration of the recording. Research staff waited outside the room and collected the readings using the EmotivPRO software from EMOTIV. Raw EEG data was exported from EmotivPRO as European Data Format (EDF) files. Data cleaning and spectral power calculation was performed using the EEGLAB open-source toolbox 2022.1 (46) in MATLAB R2019b using default parameters. Data were high passed filtered to 1 Hz to minimize the presence of non-cortical signals. All of the 14 channels filtered during the artifact removal step were interpolated using automated interpolation in EEGLAB. Spectral power calculation was completed and extracted using the Darbeliai plugin in EEGLAB (46). Waves were defined as follows: delta waves: 1 - 4 Hz, theta waves: 4 - 8 Hz, alpha waves: 8 - 13 Hz, beta waves: 13 - 32 Hz, and gamma waves: 32-60 Hz. The spectral power per waveform (alpha, beta, delta, theta, gamma) were scaled between 0 and 1 with respect to the total power across all leads and all waveforms per recording.

### Statistical Analysis

For regression models, participant mental health scores, PHQ-9 and GAD-7 scores, were normalized by log-transformed by previously adding a pseudo value of 1 and then scaled between 0 and 1.

Estimated gestational age across the antenatal period (conception to delivery) in days was scaled to between 0 and 1 and divided by the standard 40-week antenatal period and the postpartum period was scaled between 0 and 1 and then scaled to the 6 weeks postpartum divided by the standard 40-week antenatal period.

Temporal variations of the scaled and normalized mental scores were assessed with linear mixed effect models (LMMs) with the *lmer()* function in the R lme4 package (47) with GWs as fixed effect and random intercepts and random slopes at the subject level (eq.1). Models were adjusted for self-reported participant’s race, ethnicity, and use of public health insurance (i.e., Medicaid) as proxy for socioeconomic status. For both mental health assessments, we fitted models across the antenatal period only and across the entire perinatal period (pregnancy and postpartum):

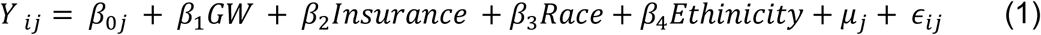

where *Y* _*ij*_ represents the normalized total score of the mental health questionnaire *i* (PHQ-9 or GAD-7 score) from individual *j*, *β*_0*j*_, and *μ*_*j*_ are the random intercept and the per individual and is the random effect per individual *j*,respectively.

Associations between the mental health scores *i* (PHQ-9 or GAD-7 score) and the overall effect of relative wave power across the antenatal and perinatal period were assessed with LMMs as before. A LLM was computed for each wave form (alpha, beta, delta, theta, gamma) as a predictor of log-centered PHQ-9 or GAD-7 and GWs and the same covariates as above for both antenatal and perinatal data. Random intercepts were included at the subject level.

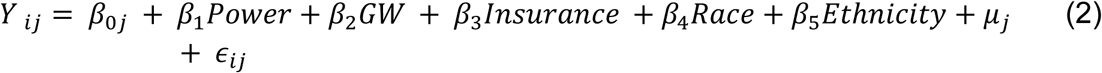

where *Power*_*j*_ represents the relative alpha, beta, delta, theta or gamma power. Participants were only included if they had a minimum of three pEEG recordings (N=46 subjects, 190 recordings).

Linear models were used to assess relationships between self-reported depression and anxiety symptoms severity (log-centered normalized) and relative power of a waveform cross-sectionally during pregnancy and postpartum (each of the five visits), correcting for race, ethnicity and insurance type PHQ-9 and GAD-7 scores were log-centered normalized, and corrected by race, ethnicity, and health insurance.

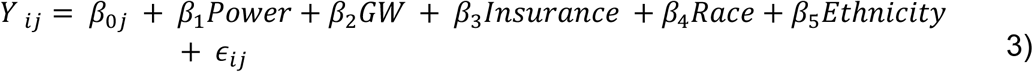

Here, *Y* _*ij*_ represents the normalized total score of the mental health questionnaire *i* (PHQ-9 or GAD-7 score) from individual *j*

We used linear models to test if early pregnancy (visit 1, <16 GWs, and visit 2, 20 GWs) spectral alpha, beta, delta, theta, and gamma power could be a prospective marker of self-reported mental health symptoms in late gestation (visit 4, ∼ 36 GWs) and postpartum (visit 5, ∼6 weeks post-delivery), linear models were used that were corrected for the same covariates

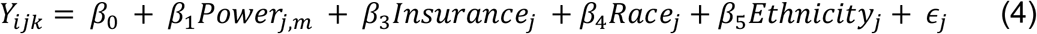

where *Y*_*ij*௞_ represents the normalized total score of the mental health questionnaire *i* (PHQ-9 or GAD-7 score) from individual *j* at visit *k* (visit 1, <16 GWs; visit 2, ∼20 GWs; visit 3, ∼28 GWs; visit 4, ∼ 36 GWs; visit 5, ∼6 GWs postdelivery).

Fisher’s exact test (48) was used to test for differences in the proportions of PHQ-9 and GAD-7 severity (normal, mild, moderate/severe) across the visits. Findings were considered significant at a p-value < 0.05 unless stated otherwise. All statistical analyses were conducted in R Studio version 024.04.2+764 (49)

## RESULTS

### Subject demographics

We analyzed a total of 209 EEG recordings from 61 different pregnant individuals, with more than 75% participants completing three or more research visits (median=4 visits). Research visits were at 13±2.1 gestational weeks (visit 1, n=49), 20.4±2.1 gestational weeks (visit 2, n=35), 26.6±1.9 gestational weeks (visit 3, n=49) 35.6±1.4 gestational weeks (visit 4, n=38), and 6.5±1.3 weeks postpartum (visit 5, n=38). Overall, the sample was composed of 45.9% Black and 14.8% Hispanic women, and 60.7% of them used public health insurance for their obstetric care (**Table 1**). The rate of moderate or severe levels of depression and anxiety per subject were 18% and 16.3%, respectively, at one or more visits in the study. Although rates of PHQ-9 and GAD-7 severity categories did not statistically differ across the perinatal period (**Figs. S1**), PHQ-9 and GAD-7 score severity did statistically vary by visit (**Figs. S2**).

**Table 1:**
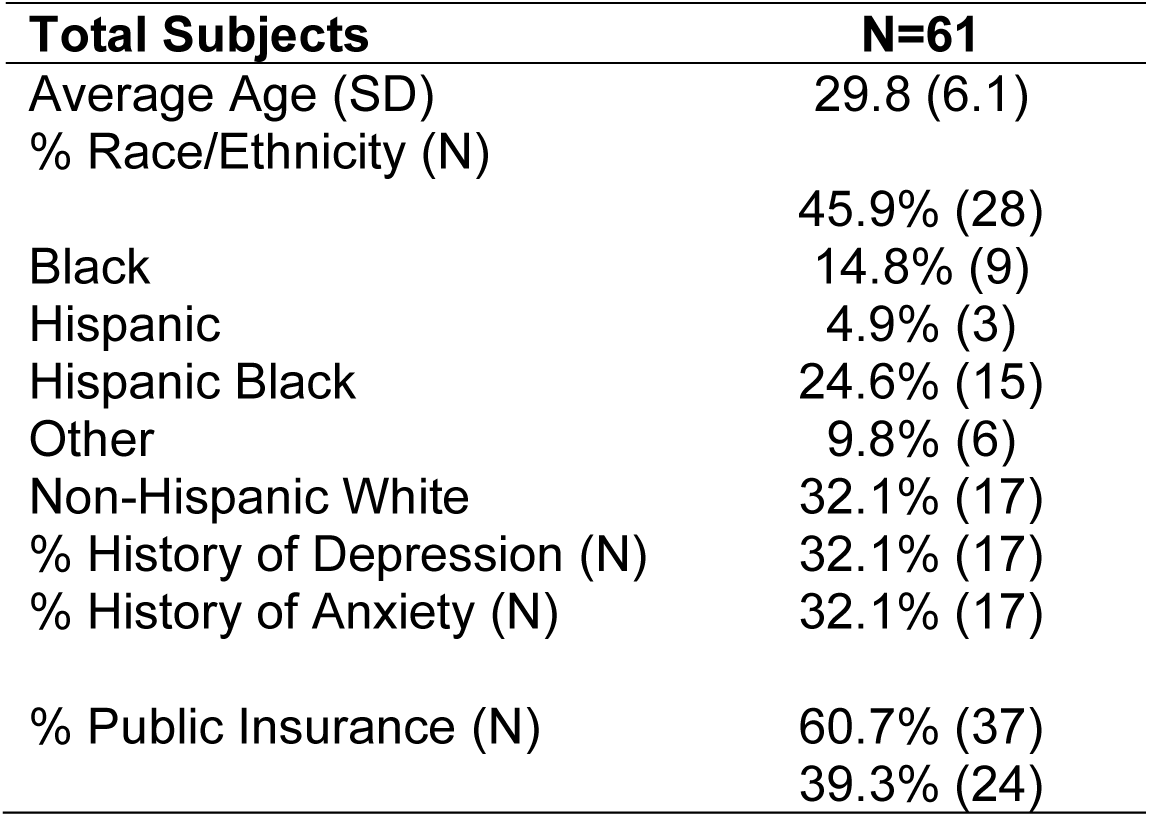
Study demographics for all subjects. History of anxiety and depression was self-reported.

### Perinatal anxiety, but not perinatal depression, across the entire perinatal period was associated with changes in brain functionality

First, we compared the associations between the temporal changes in the relative powers of alpha, beta, delta and gamma waves and mental health severity across the perinatal period (early pregnancy up to 6 weeks postpartum) using linear mixed effect models and correcting for race, ethnicity, and health insurance status. Relative delta wave power was negatively associated with perinatal anxiety symptoms but did not show a linear association with gestational age (**Fig. 2**, p<0.05). No other relative wave power across the perinatal period was significantly associated with perinatal anxiety or depression (**Table S1-2**).

**Figure 2:**
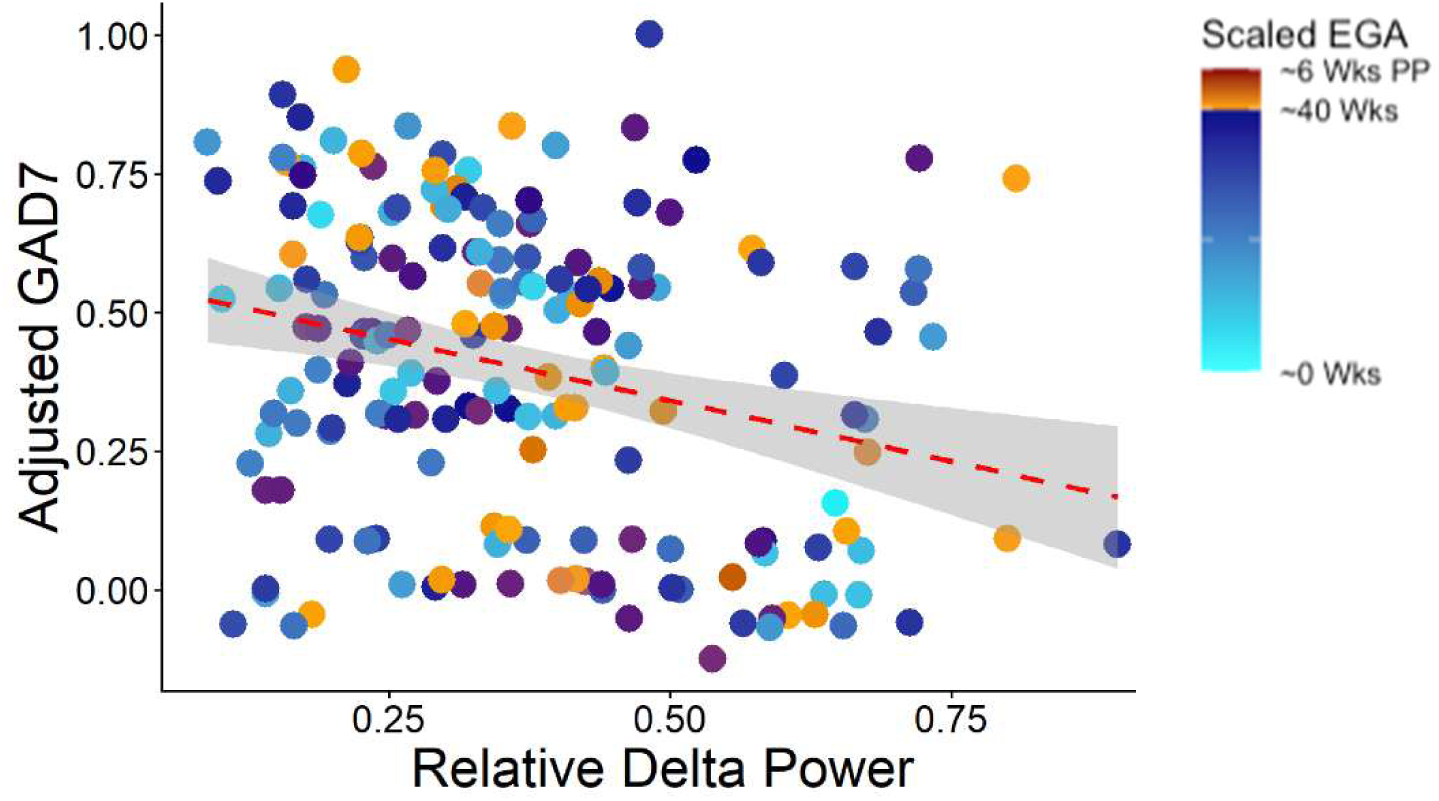
Brain waves alterations were associated with perinatal anxiety symptomatology across the entire perinatal period (early pregnancy up to 6 weeks postpartum). Perinatal delta waves were the only frequency band associated with normalized GAD7 scores (p<0.05). No frequency was associated with normalized PHQ-9 across the entire perinatal period. For visualization purposes, we plotted the residual values of the normalized total score of the mental health questionnaires *Ŷ*_*ij*_ = *Y*_*i*,*j*_ − *β*_2_*EGA* − *β*_3_*Insurance* − *β*_4_*Race* − *β*_5_*Ethnicity*) where *i* represents the questionnaire type (PHQ-9 or GAD-7) and *β*_*n*_ are the regression coefficients from the linear models.

### Brain waves were associated with self-reported mental health severity at specific perinatal time points

Next, we explored whether the relative power of alpha, beta, delta and gamma waves were associated with self-reported levels of perinatal depression and anxiety cross-sectionally during pregnancy. We observed that several frequency bands were indeed associated with depression at postpartum and with anxiety in early pregnancy and at postpartum. Relative alpha power was positively linked with self-reported anxiety levels during the second trimester (∼20 weeks) and postpartum (**Fig. 3A,** p-value <0.05; **Fig. 3B,** p-value < 0.01, respectively), but no significant association between alpha waves and PHQ-9 scores was found at any time point during the perinatal period. Relative delta power was negatively associated with both GAD-7 and PHQ-9 scores at the postpartum visit (**Fig. 3C,** p-values < 0.01; **Fig. 3D,** p-value <0.05, respectively).

**Figure 3:**
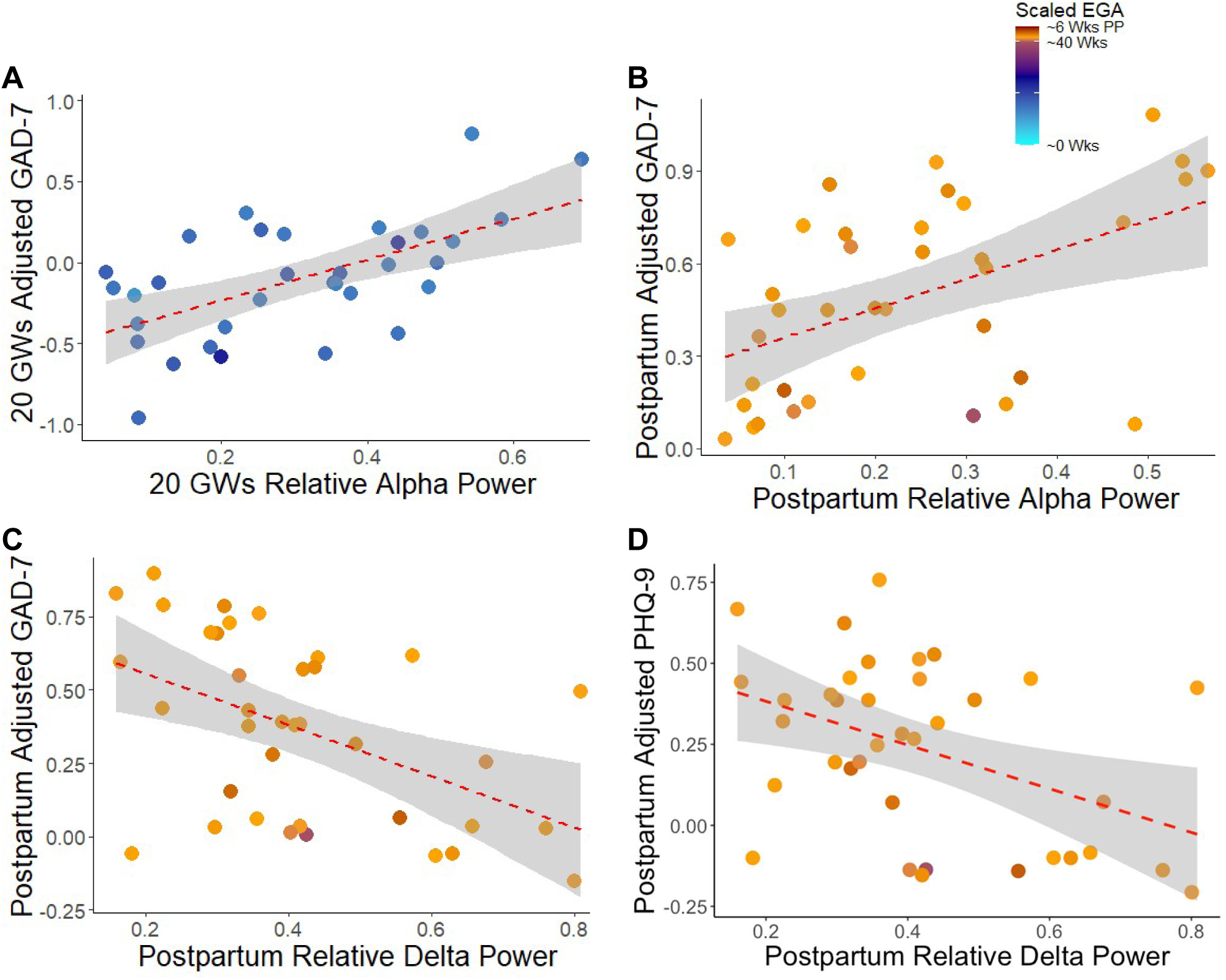
Several Relative brain wave powers were associated with depression and anxiety symptoms level at specific time points during the perinatal period. A) Relative alpha power associated with self-reported levels of anxiety (log-transformed and centered GAD-7 after removal of covariant variation) at the second trimester (∼20 GWs) (p<0.05) and (B)at postpartum (p<0.01). (C) Relative delta power negatively associated with self-reported levels of anxiety at postpartum (p<0.01) and with self-reported levels of depression (log-transformed, centered PHQ-9 after removal of covariant variation) at postpartum (p < 0.05). For visualization purposes, we plotted the residual values of the normalized total score of the mental health questionnaires (see legend of Fig. 2).

Relative theta and beta power showed no statistically significant associations with either GAD-7 or PHQ-9 scores (**Tables S3-S4,** p-value>0.05). Relative gamma power was significantly associated with self-reported anxiety levels in the postpartum (**Table S3,** p-value<0.01). In summary, alpha, delta, and gamma waves were associated with perinatal depression and anxiety during the perinatal period, yet their associations were distinct and gestationally dependent.

### Early pregnancy brain waves are prospectively associated with late pregnancy self-reported mental health severity

Finally, we investigated whether the relative power of the different brain waves earlier in pregnancy (<16 GWs, 20 GWs) were associated with the self-reported levels of depression and anxiety symptoms in late pregnancy (third trimester; 36 GWs) and postpartum. We found that the first trimester (<16 GWs) relative delta power was negatively associated with third trimester (∼36 GWs) depression and anxiety levels (**Fig. 4A-B, Tables S5 & S6**). This was mirrored at 20 GWs, where delta power was negatively associated with third trimester depression and anxiety levels (**Fig. 4C-D, Tables S9-10**). The 20 GWs alpha power was positively associated with anxiety at the third trimester and postpartum (**Figure 4E-F, Table S11**), but this association was not found for depression (**Tables S9-S10**). A significant association between third trimester GAD-7 scores and 20 GWs beta wave power was also observed (**Table S9**). However, no waveforms at 16 or 20 GWs were significantly related to postpartum depression (**Tables S10 & S12**). Taken together, brain waves in early pregnancy were associated with elevated depression and anxiety in late pregnancy and postpartum, regardless of depression and anxiety severity from other timepoints throughout the perinatal period

**Figure 4:**
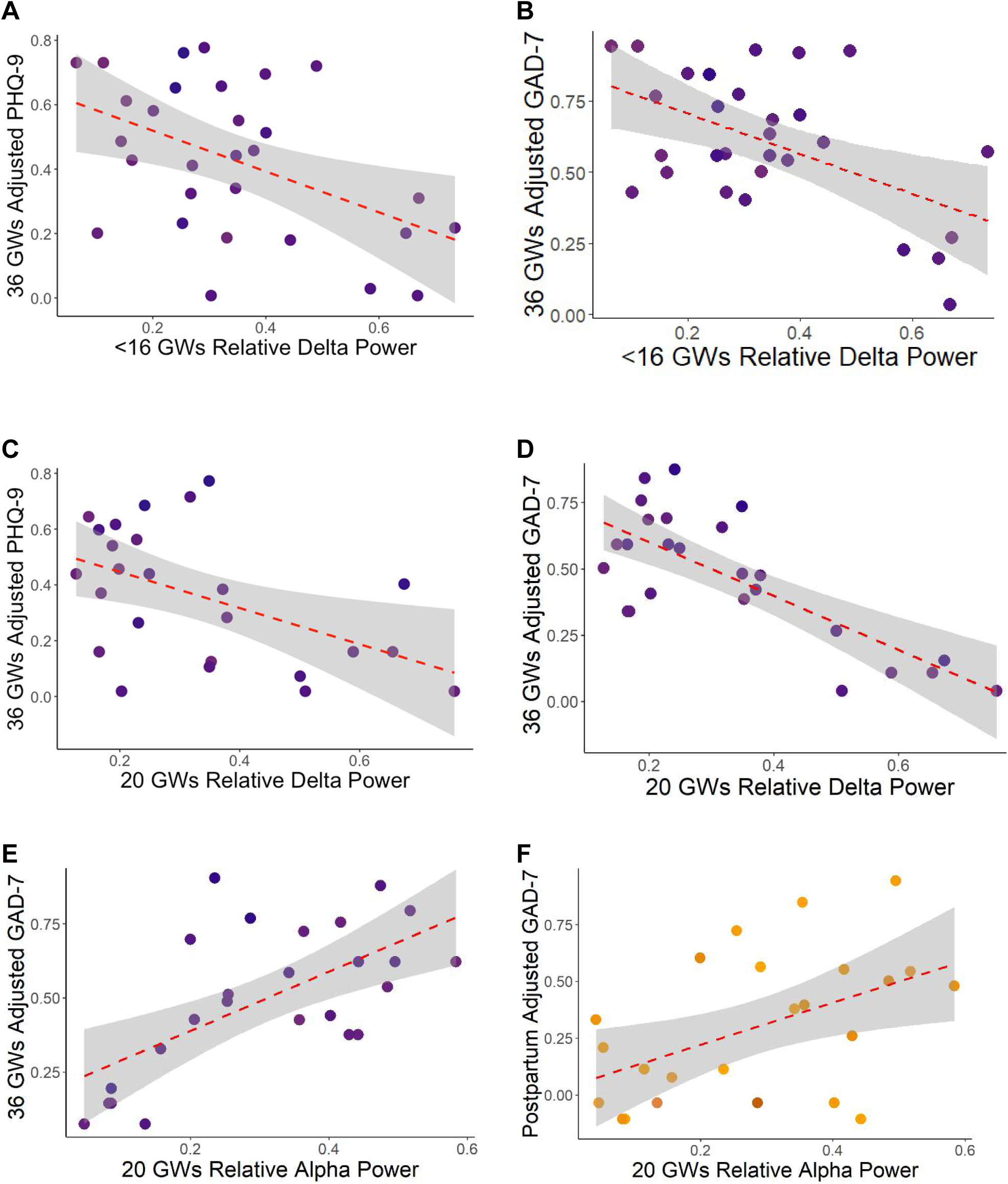
Alpha and delta relative wave power in early pregnancy were third trimester self-reported depression and anxiety severity. (A) First trimester (<16 GWs) relative delta power was negatively associated with normalized self-reported levels of depression at the third trimester (∼36 GWs) (p < 0.05) as well as (B) normalized self-reported levels of anxiety at the third trimester (∼36 GWs) (p<0.01). (C) Second trimester (20 GW) relative delta power was negatively associated with normalized self-reported levels of depression at the third trimester (∼36 GWs) (p < 0.05) as well as (D) normalized self-reported levels of anxiety at the third trimester (∼36 GWs) (p<0.001). (E) Second trimester (20 GW) relative alpha power was positively associated with normalized self-reported levels of anxiety at the third trimester (∼36 GWs) (p<0.01) and with (F) normalized self-reported levels of anxiety at the postpartum (p<0.05). For visualization purposes, we plotted the residual values of the normalized total score of the mental health questionnaires (see legend of Fig. 2).

## DISCUSSION

The perinatal period is a time of dramatic and dynamic physiological adjustments, which may impact the onset of perinatal depression and anxiety. Despite the importance of discerning the pathophysiology of perinatal mood and anxiety disorders, brain electrophysiology has never been longitudinally quantified in a perinatal population and available studies are scarce especially in the context of perinatal mental health (35,36). This study is the first to demonstrate that elevated symptoms of depression and anxiety during pregnancy and postpartum are linked to EEG-measured brain activity, specifically with alpha and delta waves (**Fig. 5**).Alpha and delta waves have been previously implicated in mental health outside of pregnancy such as major depressive disorder and generalized anxiety disorder (27,29). Alpha wave disturbances may relate to mental health associate changes in attention and relaxation while delta wave alterations may reflect deep sleep related changes(50). However, this is the first time these waves together have been linked to perinatal mood and anxiety disorders, with delta waves negatively linked with perinatal mental health severity and alpha waves positively associated with perinatal mental health severity. This may reflect sleep alterations found in perinatal depression(51) and anxiety(52) and a shift to increased restlessness or vigilance states.

**Figure 5:**
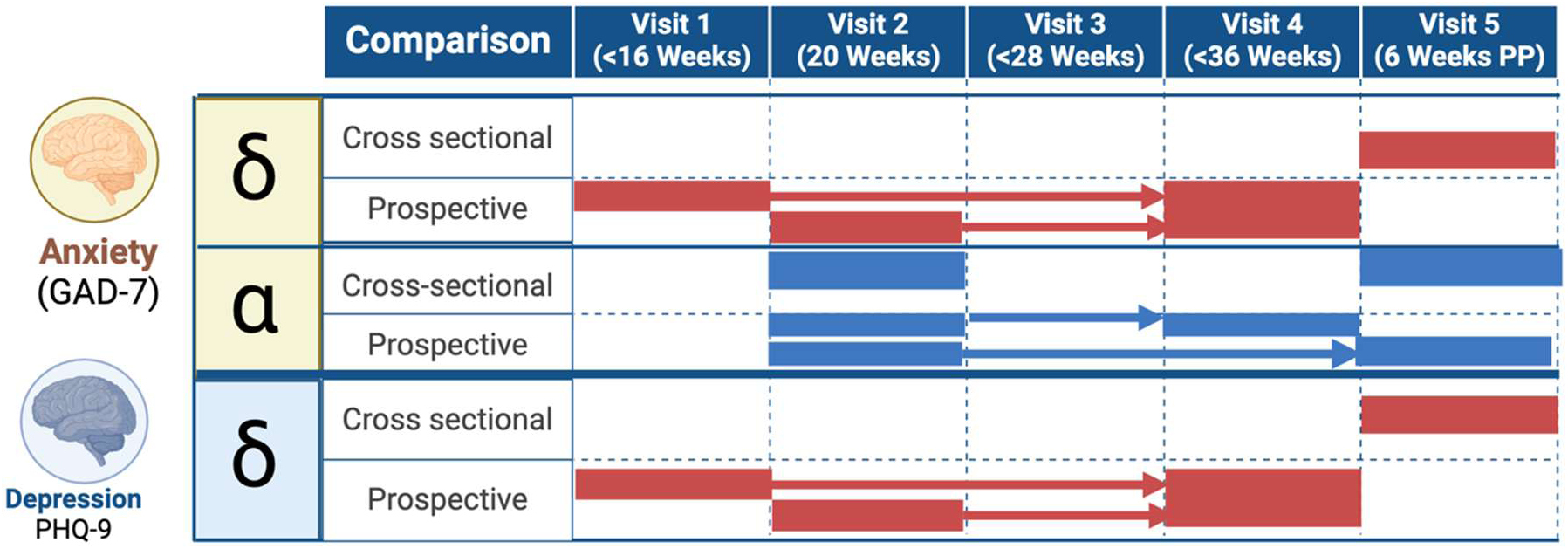
Summary of findings from cross-sectional and prospective analysis. Red indicates a negative relationship; blue indicates a positive relationship. Figure created with Biorender.

We identified a longitudinal negative association between delta waves and perinatal anxiety across the entire perinatal period (**Fig. 2**) and cross-sectionally the negative association between anxiety symptoms and delta waves was statistically significant only at postpartum (**Fig. 3**), suggesting a non-linear association between brain electrophysiology and perinatal mood disorders may not be linear across the entire perinatal period. Similarly, a non-linear association was identified with anxiety symptoms severity, but not with depression, with alpha waves positively associated at both 20 gestational weeks and postpartum. This suggests that the brain may undergo time-dependent neural adaptations throughout the perinatal period specifically at early pregnancy (i.e., ∼16 and 20 gestational weeks) and postpartum. Given that alpha waves are higher frequency waves and delta waves are lower frequency waves, this may reflect a shift towards higher frequency brain dynamics and thus a potential excitatory transition in brain electrical activity. This excitatory shift may be reminiscent of the increased alertness and vigilance found in perinatal mental health disorders. Further work must be completed to understand these potential implications to the balance of the excitation-inhibition ratio in the perinatal brain. As we expand the limited perinatal neuroimaging studies (17,35,36), we expect to better understand the pathophysiology of perinatal mental health and its difference from the physiology of health pregnancy.

In the postpartum period, our results showed a negative association between delta waves and elevated depression and anxiety symptomatology. Delta waves are instead negatively associated with perinatal mental health severity. This may underlie a potential perinatal-specific mechanism for mental health pathology. In the perinatal period, previous work by Peng and colleges assessed EEG network features in perinatal depression using recordings from 13 pregnant women with clinically diagnosed perinatal depression compared to control recordings from 29 healthy pregnant women at one timepoint in the third trimester (∼38 GWs).Based on linear discriminant analysis, they identified spatial distributions of the overall power characteristic of perinatal depression rather than specific frequency-specific waveforms. In a sample of 135 EEGs at the third trimester (>27 GWs), perinatal anxiety was positively associated with delta waves and negatively associated with beta waves has been reported to be negatively associated (36). While we did not observe these beta wave (13-32 Hz) findings, we did note slightly slower alpha wave (8-13Hz) associations with perinatal anxiety (**Fig. 3-4**) outside of the third trimester.

Of great importance for prevention, our results revealed that EEG-assessed brain waves may provide early-stage indicators and potential markers of therapeutic efficacy for future perinatal mental health conditions. We found that lower delta power in the first trimester (<16 GWs) and mid second trimester (20 GWs) was associated with higher severity of both anxiety and depression in the late third trimester (**Fig. 4**). In contrast, higher alpha power measured at 20 GWs was associated with higher anxiety in both the third trimester and the postpartum (**Fig. 4**). Schwartzmann and colleagues have shown that in non-pregnant individuals with MDD, an early increase in relative delta power and decrease in relative alpha power from baseline to two weeks was associated with greater response to cognitive behavioral therapy (53). These findings support the potential application of early pregnancy electrophysiological screening for late gestation and postpartum mental health outcomes. These EEG signatures may lead to the identification of clinically relevant quantitative markers of high-risk mental health changes early enough to prevent the onset of perinatal depression and anxiety disorders later during gestation and after delivery.

## STRENGTHS AND LIMITATIONS

To our knowledge, this study is the first to longitudinally assess brain electrical activity in relation to depression and anxiety using pEEG, from early pregnancy into the postpartum period. While other cross-sectional studies have determined the association between electroencephalogram features and prenatal mood disorders, those have been focused on late pregnancy only (14,33,35,36,54). Early pregnancy is a time of rapid, dramatic hormonal changes which may underlie the initiation of these neurological adaptations and its relationship to perinatal mental health (55,56). Our study underscores the importance of assessing brain activity early in pregnancy as it could be a novel unbiased biomarker to identify patients at high-risk of perinatal mood and anxiety disorders later in the perinatal period.

Importantly, we have increased the richness of populations included in perinatal EEG studies. Our cohort is mostly composed from low-income women of color, who have been underrepresented in mental health studies despite being disproportionally affected by them (10,57). Among other factors, he equipment typically used for EEG collection is a barrier for Black women (58) due to factors such as reduced scalp contact in those with coarse/curly hair (37). This limits the generalizability of the few existing EEG pregnancy studies to the general population. As we utilized devices whose electrode placement could be adjusted to improve contact across a variety of scalps including those with coarse/curly hair, we have been able to increase our inclusivity.

There are a multitude of critical avenues for future research opportunities. While we found some preliminary differences in brain electrical activity associated with perinatal depression and anxiety, larger follow-up longitudinal studies are required to validate and determine the robustness of our findings, and to further provide a deeper understanding of neurological processes of perinatal mental health. While the 14-lead portable EEG used in this study has greater potential to be translated into the clinic, as it is easier to use, and is more affordable compared to other research-level EEGs, it limits the spatial resolution. Therefore, to generate a more granular understanding of the role of brain electric activity in perinatal mood disorders, our results should be validated using a standard clinical or research EEG system such as a device with 128 or more electrodes (59) that can provide adequate scalp contact for individuals who may have coarse/curly hair, braids, or wigs. While we were able to interpolate any channels that were dropped during data processing and cleaning due to inadequate contact, there is a strong need for improved EEG devices that are suitable for a broader patient population. Finally, it is important to mention that our study ends after 6 weeks postdelivery. However, longitudinal up to at least one year postpartum, are necessary to determine whether the brain returns to its initial configuration prior to conception and their effect on maternal mental health.

## CONCLUSION

Maternal mental health, and mental health in general, necessitates a biological understanding of its pathophysiology. We urgently require both a more comprehensive understanding of how the brain is altered during the perinatal period and what differentiates normal perinatal physiology from pathophysiology resulting from mental health disorders such as depression and anxiety. Thus, the associations we have unveiled between delta and alpha waves and perinatal mental health symptomatology have the potential to help clinicians better identify anxiety and depression pathophysiology during the perinatal period and predict their future onset as well as understand their complex interactions since the prevalence of comorbid self-reported antenatal anxiety and mild-severe depressive symptoms is estimated to affect ∼9.5% of all pregnancies(60). We expect that our study which linked brain electrical activity and perinatal mood disorders encourages future studies in this area.

## ACKNOWLEDGEMENTS

BPB generated and conceptualized the original idea, led the planning and execution of the project and secured funding. BPB, AL, OA and MMK developed the study design. SAA, EW, AP, SP, and CC collected the experimental EEG measurements; MA and MK completed data cleaning; and SAA and MA built the computational pipeline to conduct all the analysis. BPB, AL, SAA, MA and PMM interpreted the results. MA, SAA, AL and BPB wrote the first draft of the manuscript, and all the authors critically reviewed the manuscript. MA was funded by the Neuroscience of Mental Health T32 (5T32MH067631). SAA and MA were funded by the University of Illinois Medical Scientist Training Program. This project was funded by the Yong Investigator Award from the Brain and Behavior Research Foundation and by the BIRCWH K12 Award to BPB (K12AR084225)

## DATA AVAILABILITY

All R scripts utilized in the data analysis can be found on GitHub at https://github.com/LabBea/pEEG_MentalHealth_24. Raw pEEG recordings can be made available upon request.

## SUPPLEMENTAL FIGURES

**Figure S1:**
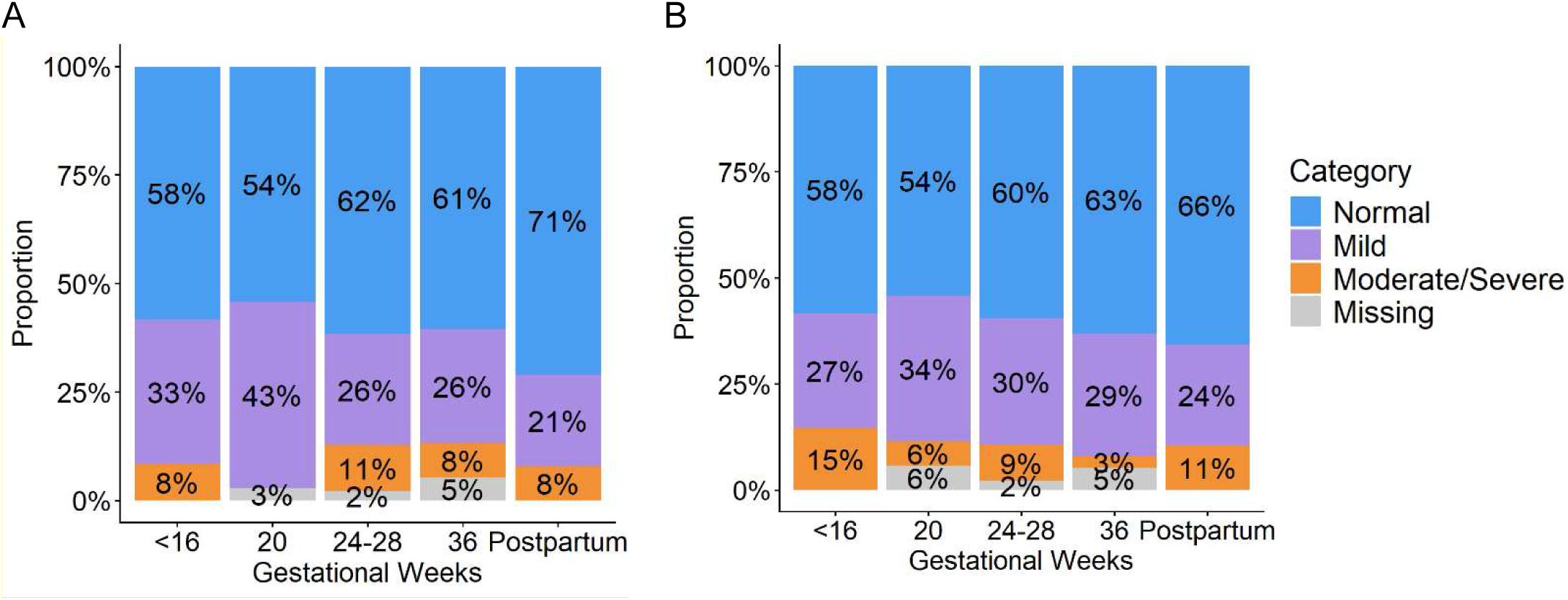
Depression and anxiety severity vary across the perinatal period. No significant differences per visit (Fisher’s exact test p-value>0.05) (A) PHQ-9 categories; Normal (0-4), Mild (5-9), and Moderate/Severe (>=10). (B) GAD-7 categories; Normal (0-4), Mild (5-9), and Moderate/Severe (>=10).

**Figure S2:**
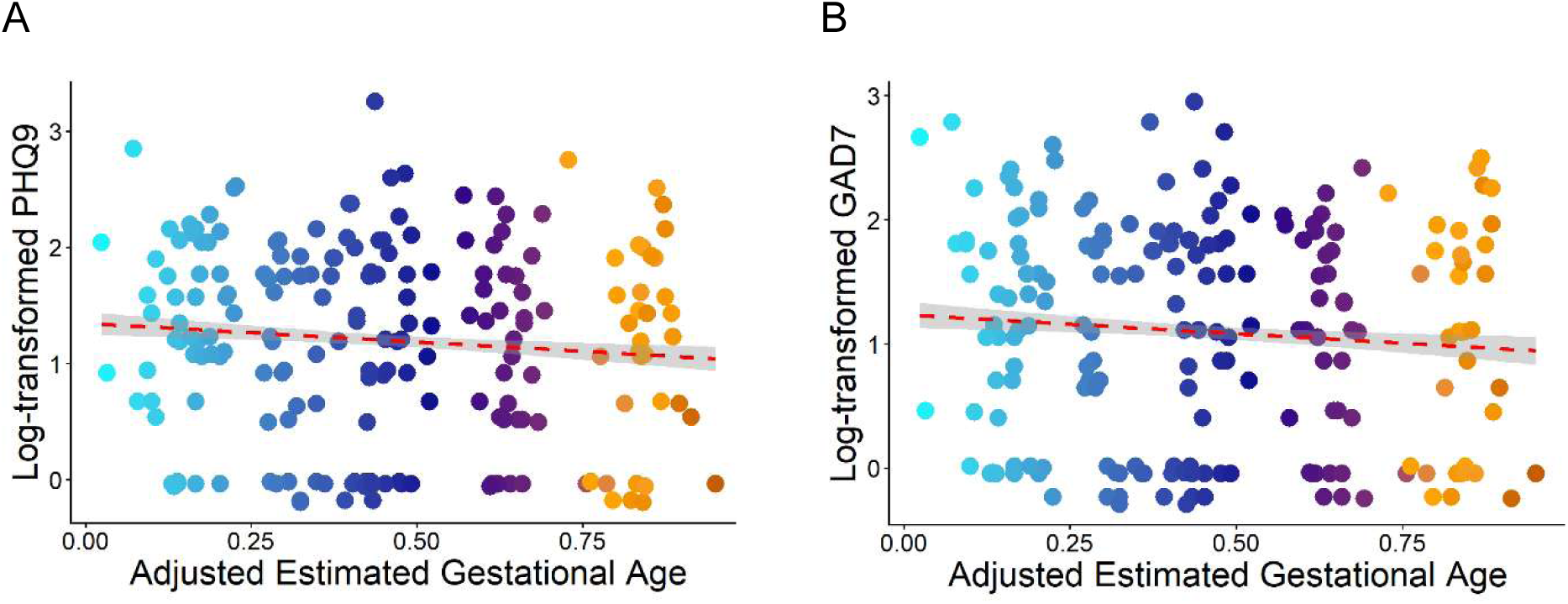
Depression and anxiety severity vary across the perinatal period and antenatal period. (A) Scaled PHQ-9 across the entire perinatal period, from early pregnancy to 6 weeks postpartum (p<0.001). (B) Scaled GAD-7 scores across the perinatal period, from early pregnancy to 6 weeks postpartum (p < 0.05).

## SUPPLEMENTAL TABLES

**Table S1: GAD-7 variations during the perinatal period.** Centered-log corrected GAD-7 scores were the outcome variable based on relative wave power from all visits. All linear mixed models were corrected by race, ethnicity, health insurance status, and gestational weeks (GWs) for that visit. Significant p-values are bolded.

**Table S2: PHQ-9 variations during the perinatal period.** Centered-log corrected PHQ-9 scores were the outcome variable based on relative wave power from all visits. All linear mixed models were corrected by race, ethnicity, health insurance status, and gestational weeks (GWs) for that visit. Significant p-values are bolded.

**Table S3: GAD-7 variations across specific periods during pregnancy and postpartum.** Centered-log GAD-7 scores were the outcome variable. All multilinear models were corrected by health insurance status and estimated gestational weeks (GWs) for that visit. 6 PP represents 6 weeks postpartum. Significant p-values are bolded.

**Table S4: PHQ-9 variations across specific periods during pregnancy and postpartum.** Centered-log PHQ-9 scores were the outcome variable. All multilinear models were corrected by health insurance status and gestational weeks (GWs) for that visit. 6 PP represents 6 weeks postpartum. Significant p-values are bolded.

**Table S5: Third trimester centered-log GAD-7 scores were associated with the first trimester spectral power.** All models were corrected by race, ethnicity, health insurance status, and gestational weeks (GWs) for that visit. Significant p-values are bolded.

**Table S6: Third trimester centered-log PHQ-9 scores were associated with the first trimester spectral power.** All models were corrected by race, ethnicity, health insurance status, and gestational weeks (GWs) for that visit. Significant p-values are bolded.

**Table S7: Postpartum centered-log GAD-7 scores were associated with the first trimester spectral power.** All models were corrected by race, ethnicity, health insurance status, and gestational weeks (GWs) for that visit. Significant p-values are bolded.

**Table S8: Postpartum centered-log PHQ-9 scores were associated with the first trimester spectral power.** Postpartum centered-log PHQ-9 scores were the outcome variable based on the first trimester spectral power. All models were corrected by race, ethnicity, health insurance status, and gestational weeks (GWs) for that visit. Significant p-values are bolded.

**Table S9: 20 GWs Spectral Power & Third Trimester GAD-7.** Third trimester centered-log GAD-7 scores were the outcome variable based on the 20 GW spectral power. All models were corrected by race, ethnicity, health insurance status, and gestational weeks (GWs) for that visit. Significant p-values are bolded.

**Table S10: 20 GWs Spectral Power & Third Trimester PHQ-9.** Third trimester centered-log PHQ-9 scores were the outcome variable based on the 20 GWs spectral power. All models were corrected by race, ethnicity, health insurance status, and gestational weeks (GWs) for that visit. Significant p-values are bolded.

**Table S11: 20 GWs Spectral Power & Postpartum GAD-7.** Postpartum centered-log GAD-7 scores were the outcome variable based on the 20 GWs spectral power. All models were corrected by race, ethnicity, health insurance status, and gestational weeks (GWs) for that visit. Significant p-values are bolded.

**Table S12: 20 GWs Spectral Power & Postpartum PHQ-9.** Postpartum centered-log PHQ-9 scores were the outcome variable based on the 20 GWs spectral power. All models were corrected by race, ethnicity, health insurance status, and gestational weeks (GWs) for that visit. Significant p-values are bolded.

